# Associations of Long-term Sleep Duration Trajectories with Overall and Cause-Specific Mortality Among Middle-to-older Aged Black and White Adults

**DOI:** 10.1101/2024.05.23.24307845

**Authors:** Kelsie M. Full, Hui Shi, Loren Lipworth, Lawrence T. Dauer, Michael T. Mumma, Qian Xiao

## Abstract

**Background:** Both short and long sleep durations are adversely associated with numerous chronic conditions, including cardiovascular disease (CVD), diabetes, hypertension, and mortality. The American Academy of Sleep Medicine recommends adults in the United States sleep at least 7 hours and less than 9 hours per night to maintain optimal health. It remains unclear how sleep duration trajectories over time are associated with mortality.

**Methods:** This observational cohort study includes 46,928 Black and White adults (mean age: 53 ± 9 years) who enrolled in the Southern Community Cohort Study between 2002–2009 and completed a follow-up survey in 2008-2013. Participants were categorized into nine sleep duration trajectory categories based on the reported average sleep duration between study enrollment and at follow-up. Participant vital status and date and cause of death were ascertained via linkage to the National Death Index through 2022. Cox regression analysis was performed to estimate hazard ratios (HR) and 95% confidence intervals (CI) for the association between sleep duration trajectory and all-cause and cause-specific mortality (CVD, cancer, and neurodegenerative disease) after adjustment for sociodemographic characteristics, health behaviors, and clinical factors.

**Results:** During a median 12.6 years of follow-up, we documented 13,579 deaths, including 4,135 from CVD, 3,067 from cancer, and 544 from neurodegenerative diseases. Compared to the optimal sleep duration trajectory (maintaining 7-9 hours), all sub-optimal trajectories were associated with significant 6 to 33% greater risk of all-cause mortality in fully adjusted models. Compared to the optimal sleep trajectory, three of the sub-optimal trajectories were associated with increased CVD mortality, with HRs ranging from 1.20 to 1.34. The short-long trajectory was associated with the greatest risk of all-cause mortality (HR:1.33; 95%CI: 1.21, 1.46) and the long-short trajectory was associated with the greatest CVD mortality risk (HR:1.34; 95%CI: 1.10, 1.65). The healthy-long trajectory was associated with the greatest risk of cancer mortality (HR: 1.19; 95%CI:1.00, 1.41). None of the sub-optimal trajectories was associated with neurodegenerative disease mortality.

**Conclusions:** Suboptimal sleep duration trajectories were associated with increased risk of all-cause mortality as well as CVD mortality. Findings highlight the importance of maintaining healthy sleep duration throughout midlife to reduce mortality risk.

## Introduction

The American Academy of Sleep Medicine, National Sleep Foundation (NSF), and American Heart Association recommend adults sleep at least 7 hours^1^ and less than 9 hours^2^ per night to maintain optimal overall health and cardiovascular health.^3^ Currently, only 30-45% of adults in the United States (US) are meeting these recommendations.^4^ In a recent study conducted in the National Health and Nutrition Examination Surveys (NHANES), 35% of participants reported sleeping ≤6 hours per night.^5^ Compared to other race-ethnicity groups, the highest prevalence of short sleep duration in the NHANES study was reported among Non-Hispanic Black adults,^5^ a finding that is consistent with evidence supporting the well-documented sleep disparities across racial/ethnic groups.^6,7^ Sex and gender differences in sleep health have also been observed, suggesting that men and women sleep differently.^8^ Despite these disparities, the age-adjusted prevalence of reporting difficulty sleeping increased from 2005 to 2018 in the overall population, including among both men and women and in all race/ethnicity groups,^5^ highlighting the need for public health strategies to address poor sleep health at the population level.

Abnormal sleep duration has been associated with numerous adverse health outcomes including cardiovascular disease (CVD),^9–11^ type 2 diabetes,^12^ cancer, and mortality.^13,14^ A U-shaped relationship between sleep duration and mortality has been consistently observed, indicating that both short and long sleep durations increase the risk of death.^13–15^ In a meta-analysis of 28 studies, short, and particularly long sleep duration was associated with a 4-24% greater risk of all-cause mortality.^14^ Subgroup analyses revealed the association between long sleep duration and mortality was stronger in women than in men.^14^ In the UK biobank cohort among 407,500 adults, self-reported short (≤5 hours) and long (≥9 hours) sleep duration were both associated with higher risk of all-cause mortality (HR = 1.25, 95% CI: 1.16–1.34 and HR = 1.30, 95% CI: 1.22–1.38, respectively), as well as CVD mortality (HR = 1.27, 95% CI: 1.09–1.49 and HR = 1.32, 95% CI: 1.16–1.50, respectively).^13^ Long sleep duration was also associated with a higher risk of cancer mortality (HR = 1.19, 95% CI: 1.10–1.30).^13^ While this evidence supports a link between abnormal sleep duration and mortality, most of the included studies only measured sleep duration at one time point. Less is known about how changes in sleep duration over time may be related to mortality risk. The existing evidence is further limited by predominately homogenous non-Hispanic White samples or studies that lack sufficient power to look at important subgroup differences.

To fill this important evidence gap, we investigated the association of sleep duration trajectories over time with risk of all-cause and cause-specific mortality in the Southern Community Cohort Study (SCCS), a large ongoing US cohort study of primarily low-income middle-to-older aged Black and White adults. In this study we leveraged repeat measures of habitual sleep duration within a cohort that is large enough to investigate if associations differ by sex, race, and socioeconomic status (SES). We hypothesize that all sleep duration trajectories that differ from maintaining an optimal healthy sleep duration of 7-9 hours per night are associated with higher risk of mortality in both Black and White adults and across sex and SES subgroups.

## Methods

### Study population

Nearly 86,000 adults, age 40-79 years, from twelve Southeastern US states (Alabama, Arkansas, Florida, Georgia, Kentucky, Louisiana, Mississippi, North Carolina, South Carolina, Tennessee, Virginia, and West Virginia) were recruited and enrolled in the SCCS from 2002-2009.^16^ Eligible adults were recruited primarily from Community Health Center partners that primarily served uninsured and underinsured populations. A detailed description of SCCS methods has been previously published^16–18^ and is available on the study website (http://www.southerncommunitystudy.org/). SCCS participants provided informed consent and all study protocols were approved by institutional review boards at Vanderbilt University Medical Center and Meharry Medical College.

At baseline, SCCS participants completed a questionnaire reporting information on sociodemographic characteristics, health behaviors, including sleep duration, and health and disease history, as well as an 89-item food frequency questionnaire. In 2008-2013, SCCS participants were asked to complete a follow-up survey and provide updated information on health behaviors, including sleep duration, and health status. A total of 50,644 Black and White participants completed the follow-up survey. From the sample that completed the follow-up survey, we further excluded those who were missing information on sleep duration at baseline or follow-up (N=3,679), and those who were not followed past the follow-up survey (N=37). The final analytic sample consisted of 46,928 participants (Supplemental Figure 1).

### Mortality

Participant vital status, as well as date and cause of death, was ascertained via linkage of the cohort to the National Death Index through December 31, 2022. Cause-specific categories of death were classified as CVD (National Death Index Code (NDI) 053), cancer (NDI 019), and neurodegenerative disease (combined Parkinson’s disease NDI 051, Alzheimer’s disease NDI 052, and codes for dementia).

### Sleep Trajectories

At SCCS study baseline, participants were asked “How many hours do you typically sleep in a 24-hour period?” reporting weekdays and weekends separately. Baseline sleep duration was then calculated as the weighted average of weekday and weekend sleep durations ((weekday duration*5+weekend duration*2)/7). In the follow-up survey, participants responded to the question “In a 24-hour period, how many hours do you typically spend sleeping?” At follow-up no distinction was made between weekday and weekend sleep duration. For this analysis, sleep duration was categorized as short (<7 hours), healthy (7-9 hours) and long (>9 hours) based on the current sleep duration recommendations.^19^ Next, we defined nine sleep trajectories based on the change in sleep duration category between baseline and follow-up survey: healthy-healthy (optimal), healthy-short, healthy-long, short-healthy, short-short (stable short), short-long, long-healthy, long-short, and long-long (stable long). For all analyses, the optimal healthy-healthy category served as the reference group.

### Covariates

Relevant covariates were obtained from the SCCS baseline questionnaire. Demographic variables included age at enrollment, sex, and race (White vs. Black). Individual SES indicators included education (categorized as ≤some high school or high school graduate or higher), marital status (married/living as married with a partner vs. separated/divorced/widowed/single), household income (≥$15,000 vs. <$15,000), and employment status (unemployed vs. employed). Health behaviors included smoking status (current smoker vs. non-current smoker), alcohol intake (≤1 drink per day, >1 drink per day), diet quality, physical activity, and sedentary time. Diet quality was estimated with the Healthy Eating Index (HEI 2010),^20^ which was calculated by linking the food frequency questionnaire data with MyPyramid Equivalents Database. HEI 2010 scores are calculated (0–100 points) based on the *Dietary Guidelines for Americans*.^20^ Total moderate physical activity (MET-hours) was estimated by summing the energy expenditure of moderate occupation and sport related activity, and then categorized into quartiles. Sedentary time was estimated by summing the reported total hours spent in sitting activities, and then categorized into quartiles. Body mass index (BMI) was calculated as weight in kilograms divided by height in meters squared. Health conditions included a self-reported history of physician diagnosis of hypertension, type 2 diabetes, hyperlipidemia, heart attack or myocardial infarction (MI).

### Statistical Analyses

Participants’ baseline demographic, socioeconomic, behavioral, and health characteristics were described by sleep duration trajectory category. Descriptive statistics are presented as mean (standard deviation (SD)) for normally distributed continuous variables, median (interquartile range, IQR) for continuous variables with skewed distributions, and frequency (percentage) for categorical variables. Due to the large sample size, we did not present p-values for differences in descriptive statistics.

Kaplan-Meier plots were used to visualize the associations between sleep trajectory and all-cause mortality and to confirm the proportional hazard assumption was met. Plots were adjusted for age, sex, and race.

Survival time was calculated as the time between the date of first follow-up survey and the date of death or December 31, 2022. To examine the association between sleep duration trajectory and all-cause mortality we ran a series of adjusted Cox proportional hazard regression models. Next, Fine-Gray competing risks regression models were used for the CVD-, cancer-, and neurodegenerative disease-specific mortality outcomes. Four regression models were fitted sequentially for each mortality outcome variable. Model 1 was adjusted for age, sex, race. Model 2 added adjustment for individual SES variables, including education, marital status, household income, and employment status. In our primary model, model 3, we further adjusted for health behaviors, including: smoking status, alcohol intake, diet quality, moderate physical activity, and sedentary time. The fully adjusted model (model 4) included adjustment for relevant health conditions that may be potential mediators, including BMI, hypertension, diabetes, and high cholesterol.

We conducted several sensitivity analyses. We tested for effect modification by sex, race, and SES indicators (education and household income). The cutoff points for education and income were chosen to create roughly equal sample size in subgroups to preserve statistical power. These preliminary analyses showed evidence of an interaction between sleep trajectory*race and sleep trajectory*income, thus we reran analyses separately in White and Black participants for all-cause and disease-specific mortality outcomes. We repeated the CVD-specific mortality models after excluding participants who reported an MI at baseline or first follow-up time (N=5,373). We also repeated all-cause and cause-specific models after excluding participants who died within the first 2 years of follow-up to address the potential for reverse causation. To address the issue of missing data, we employed Multiple Imputation by Chained Equations (MICE) to impute missing values of covariates. The Cox proportional hazards regression models were repeatedly run using 20 imputed datasets to examine the associated between sleep trajectory and all-cause mortality.

All analyses were performed using R statistical software version 4.3.1.^21^

## Results

The mean age of participants at baseline was 53.0 ± 8.8 years, with 65% of the sample identifying as women and 63% identifying as Black or African American (**Supplemental Table 1**). Approximately one half (49%) of participants reported an average household income <$15,000 a year, 34% reported being a current smoker, and 56% reported having hypertension. The average sleep duration reported at enrollment was 7 hours ± 1.9 per night.

Participant baseline characteristics according to sleep trajectory category are presented in **Table 1**. Just over 33% of participants maintained an optimal 5-year sleep duration trajectory that stayed within the recommended 7-9 hours per night. Participants with an optimal trajectory were older and more likely to identify as White compared to participants in all other trajectories. Participants who maintained a stable short trajectory (short-short) or transitioned to short sleep durations (healthy-short) were more likely to be women. Participants who maintained a stable long trajectory (long-long) were more likely to be unemployed compared to all other trajectories. Participants who transitioned from short to long sleep durations (short-long) were more likely to have hypertension, diabetes, high cholesterol, and history of a previous heart attack or stroke than all other groups.

**Table 1.**
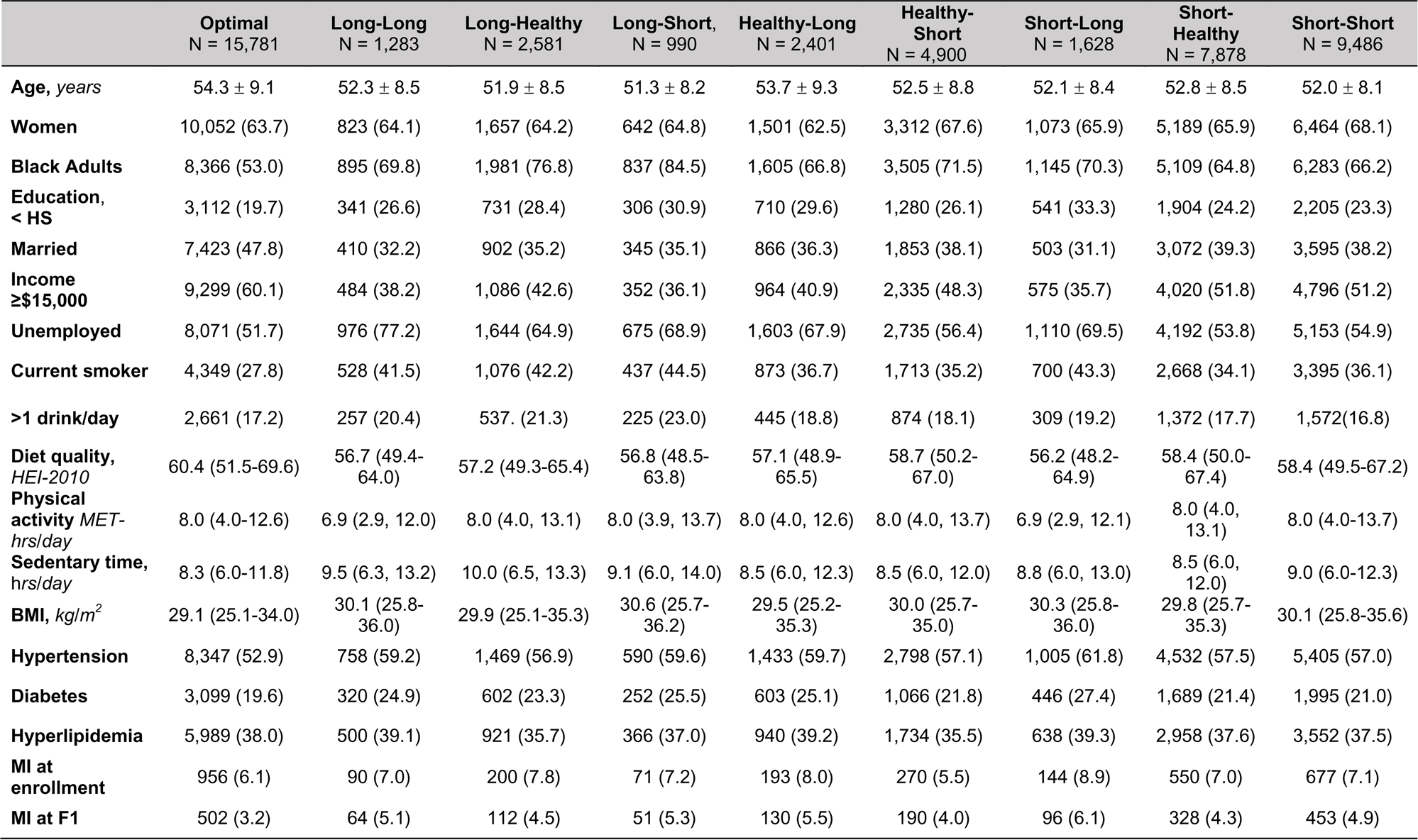
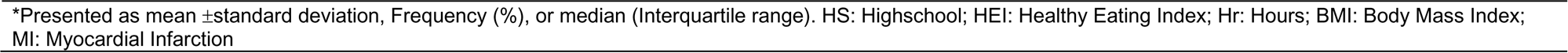
SCCS Participant Characteristics Stratified by Sleep Duration Trajectory (n=46,928)

|  | Optimal<br>N = 15,781 | Long-Long<br>N = 1,283 | Long-Healthy<br>N = 2,581 | Long-Short,<br>N = 990 | Healthy-Long<br>N = 2,401 | Healthy-Short<br>N = 4,900 | Short-Long<br>N = 1,628 | Short-Healthy<br>N = 7,878 | Short-Short<br>N = 9,486 |
| --- | --- | --- | --- | --- | --- | --- | --- | --- | --- |
| **Age, years** | 54.3 ± 9.1 | 52.3 ± 8.5 | 51.9 ± 8.5 | 51.3 ± 8.2 | 53.7 ± 9.3 | 52.5 ± 8.8 | 52.1 ± 8.4 | 52.8 ± 8.5 | 52.0 ± 8.1 |
| **Women** | 10,052 (63.7) | 823 (64.1) | 1,657 (64.2) | 642 (64.8) | 1,501 (62.5) | 3,312 (67.6) | 1,073 (65.9) | 5,189 (65.9) | 6,464 (68.1) |
| **Black Adults** | 8,366 (53.0) | 895 (69.8) | 1,981 (76.8) | 837 (84.5) | 1,605 (66.8) | 3,505 (71.5) | 1,145 (70.3) | 5,109 (64.8) | 6,283 (66.2) |
| **Education,**<br>**< HS** | 3,112 (19.7) | 341 (26.6) | 731 (28.4) | 306 (30.9) | 710 (29.6) | 1,280 (26.1) | 541 (33.3) | 1,904 (24.2) | 2,205 (23.3) |
| **Married** | 7,423 (47.8) | 410 (32.2) | 902 (35.2) | 345 (35.1) | 866 (36.3) | 1,853 (38.1) | 503 (31.1) | 3,072 (39.3) | 3,595 (38.2) |
| **Income**<br>**≥\$15,000** | 9,299 (60.1) | 484 (38.2) | 1,086 (42.6) | 352 (36.1) | 964 (40.9) | 2,335 (48.3) | 575 (35.7) | 4,020 (51.8) | 4,796 (51.2) |
| **Unemployed** | 8,071 (51.7) | 976 (77.2) | 1,644 (64.9) | 675 (68.9) | 1,603 (67.9) | 2,735 (56.4) | 1,110 (69.5) | 4,192 (53.8) | 5,153 (54.9) |
| **Current smoker** | 4,349 (27.8) | 528 (41.5) | 1,076 (42.2) | 437 (44.5) | 873 (36.7) | 1,713 (35.2) | 700 (43.3) | 2,668 (34.1) | 3,395 (36.1) |
| **>1 drink/day** | 2,661 (17.2) | 257 (20.4) | 537. (21.3) | 225 (23.0) | 445 (18.8) | 874 (18.1) | 309 (19.2) | 1,372 (17.7) | 1,572(16.8) |
| **Diet quality,**<br>**HEI-2010** | 60.4 (51.5-69.6) | 56.7 (49.4-64.0) | 57.2 (49.3-65.4) | 56.8 (48.5-63.8) | 57.1 (48.9-65.5) | 58.7 (50.2-67.0) | 56.2 (48.2-64.9) | 58.4 (50.0-67.4) | 58.4 (49.5-67.2) |
| **Physical**<br>**activity MET-**<br>**hrs/day** | 8.0 (4.0-12.6) | 6.9 (2.9, 12.0) | 8.0 (4.0, 13.1) | 8.0 (3.9, 13.7) | 8.0 (4.0, 12.6) | 8.0 (4.0, 13.7) | 6.9 (2.9, 12.1) | 8.0 (4.0, 13.1) | 8.0 (4.0-13.7) |
| **Sedentary time,**<br>**hrs/day** | 8.3 (6.0-11.8) | 9.5 (6.3, 13.2) | 10.0 (6.5, 13.3) | 9.1 (6.0, 14.0) | 8.5 (6.0, 12.3) | 8.5 (6.0, 12.0) | 8.8 (6.0, 13.0) | 8.5 (6.0, 12.0) | 9.0 (6.0-12.3) |
| **BMI, kg/m<sup>2</sup>** | 29.1 (25.1-34.0) | 30.1 (25.8-36.0) | 29.9 (25.1-35.3) | 30.6 (25.7-36.2) | 29.5 (25.2-35.3) | 30.0 (25.7-35.0) | 30.3 (25.8-36.0) | 29.8 (25.7-35.3) | 30.1 (25.8-35.6) |
| **Hypertension** | 8,347 (52.9) | 758 (59.2) | 1,469 (56.9) | 590 (59.6) | 1,433 (59.7) | 2,798 (57.1) | 1,005 (61.8) | 4,532 (57.5) | 5,405 (57.0) |
| **Diabetes** | 3,099 (19.6) | 320 (24.9) | 602 (23.3) | 252 (25.5) | 603 (25.1) | 1,066 (21.8) | 446 (27.4) | 1,689 (21.4) | 1,995 (21.0) |
| **Hyperlipidemia** | 5,989 (38.0) | 500 (39.1) | 921 (35.7) | 366 (37.0) | 940 (39.2) | 1,734 (35.5) | 638 (39.3) | 2,958 (37.6) | 3,552 (37.5) |
| **MI at**<br>**enrollment** | 956 (6.1) | 90 (7.0) | 200 (7.8) | 71 (7.2) | 193 (8.0) | 270 (5.5) | 144 (8.9) | 550 (7.0) | 677 (7.1) |
| **MI at F1** | 502 (3.2) | 64 (5.1) | 112 (4.5) | 51 (5.3) | 130 (5.5) | 190 (4.0) | 96 (6.1) | 328 (4.3) | 453 (4.9) |

\*Presented as mean ± standard deviation, Frequency (%), or median (Interquartile range). HS: Highschool; HEI: Healthy Eating Index; Hr: Hours; BMI: Body Mass Index; MI: Myocardial Infarction

### All-Cause Mortality

Over a median follow-up of 12.6 years (IQR: 11 to 13 years), there were 13,579 deaths, including 4,135 deaths from CVD, 3,067 from cancer, and 544 from neurodegenerative disease. **Figure 1** displays the Kaplan Meir survival curves of all-cause mortality by sleep trajectory adjusted for age, sex, and race. The risk of mortality over the 12 years of follow-up differed significantly between the optimal sleep trajectory and all other suboptimal trajectories (p<0.001).

**Figure 1.**
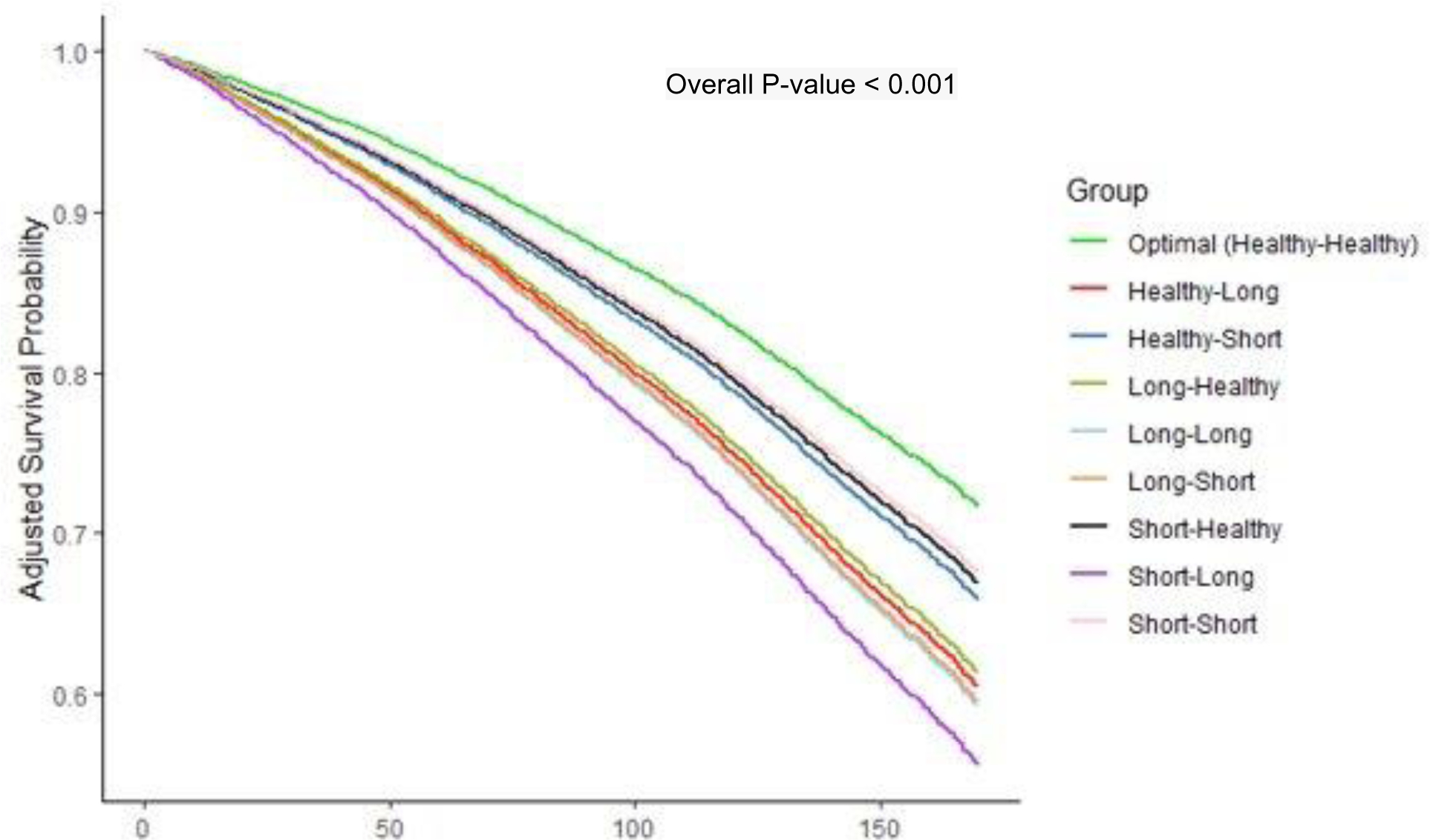
Adjusted survival curves for all-cause mortality by sleep trajectory. Adjusted for age, sex, and race.

Compared to those with an optimal sleep trajectory, participants in every other sleep trajectory category had a greater risk of all-cause mortality (**Table 2**). After adjustment for demographics, socioeconomic factors, and health behaviors (Model 3), all suboptimal sleep duration trajectories were associated with a HR for mortality ranging from 1.09 to 1.40, with the greatest risk observed among the short-long trajectory (HR: 1.40, CI: 1.28, 1.53). In fully adjusted models, with adjustment for clinical factors including BMI, hypertension, diabetes, and high cholesterol, all associations persisted, and suboptimal sleep duration trajectories were associated with a 6% (short-short; HR: 1.06, CI:1.01, 1.12) to 33% (short-long; HR: 1.33, CI: 1.21, 1.46) greater risk of mortality compared to the optimal sleep trajectory.

**Table 2.** Sleep trajectories and All-Cause Mortality in the Southern Community Cohort Study (N=46,928)

|  | N | Events | Model 1:<br>HR (95% CI)<sup>a</sup> | Model 2:<br>HR (95% CI)<sup>b</sup> | Model 3:<br>HR (95% CI)<sup>c</sup> | Model 4:<br>HR (95% CI)<sup>d</sup> |
| --- | --- | --- | --- | --- | --- | --- |
| Optimal (Healthy-Healthy) | 15,781 | 4,197 | — | — | — | — |
| Long-Long | 1,283 | 457 | 1.62 (1.47, 1.78) | 1.32 (1.20, 1.46) | 1.31 (1.18, 1.46) | 1.27 (1.15, 1.41) |
| Long-Healthy | 2,581 | 849 | 1.51 (1.40, 1.62) | 1.30 (1.20, 1.40) | 1.25 (1.15, 1.36) | 1.21 (1.11, 1.31) |
| Long-Short | 990 | 335 | 1.61 (1.44, 1.80) | 1.33 (1.19, 1.49) | 1.26 (1.11, 1.42) | 1.22 (1.08, 1.38) |
| Healthy-Long | 2,401 | 862 | 1.56 (1.45, 1.67) | 1.33 (1.23, 1.43) | 1.28 (1.18, 1.38) | 1.23 (1.13, 1.33) |
| Healthy-Short | 4,900 | 1,439 | 1.27 (1.20, 1.35) | 1.16 (1.09, 1.23) | 1.14 (1.07, 1.22) | 1.12 (1.05, 1.19) |
| Short-Long | 1,628 | 608 | 1.83 (1.68, 2.00) | 1.48 (1.35, 1.61) | 1.40 (1.28, 1.53) | 1.33 (1.21, 1.46) |
| Short-Healthy | 7,878 | 2,281 | 1.22 (1.16, 1.29) | 1.15 (1.09, 1.21) | 1.13 (1.07, 1.19) | 1.11 (1.05, 1.17) |
| Short-Short | 9,486 | 2,551 | 1.19 (1.13, 1.25) | 1.12 (1.06, 1.18) | 1.09 (1.03, 1.15) | 1.06 (1.01, 1.12) |

<sup>1</sup>HR = Hazard Ratio, CI = Confidence Interval
<sup>a</sup>Model 1: adjusted for age, sex, race;
<sup>b</sup>Model 2: model 1 + adjustment for education, marital status, household income, employment status;
Model 3: model 2 + adjustment for smoking status, alcohol intake, HEI-2010, total moderate activity, sedentary time;
Model 4: model 3 + adjustment for BMI, hypertension, diabetes, high cholesterol

### Cause-Specific Mortality

Compared to participants who maintained an optimal sleep trajectory, almost all other sleep trajectories were associated with greater risk of CVD mortality (**Table 3**). In the model adjusted for demographics, socioeconomic factors, and health behaviors (model 3) the risk for CVD mortality ranged from an HR of 1.12 to 1.40 with participants with the long-short trajectory at the greatest risk (HR:1.40; CI:1.14, 1.71). The observed associations persisted after adjustment for traditional CVD risk factors, including BMI, hypertension, diabetes, and high cholesterol, for the long-healthy (HR:1.20; CI:1.03, 1.38), long-short (HR:1.34; CI:1.10, 1.65), and short-long trajectories (HR:1.23; CI:1.04, 1.46).

**Table 3.**
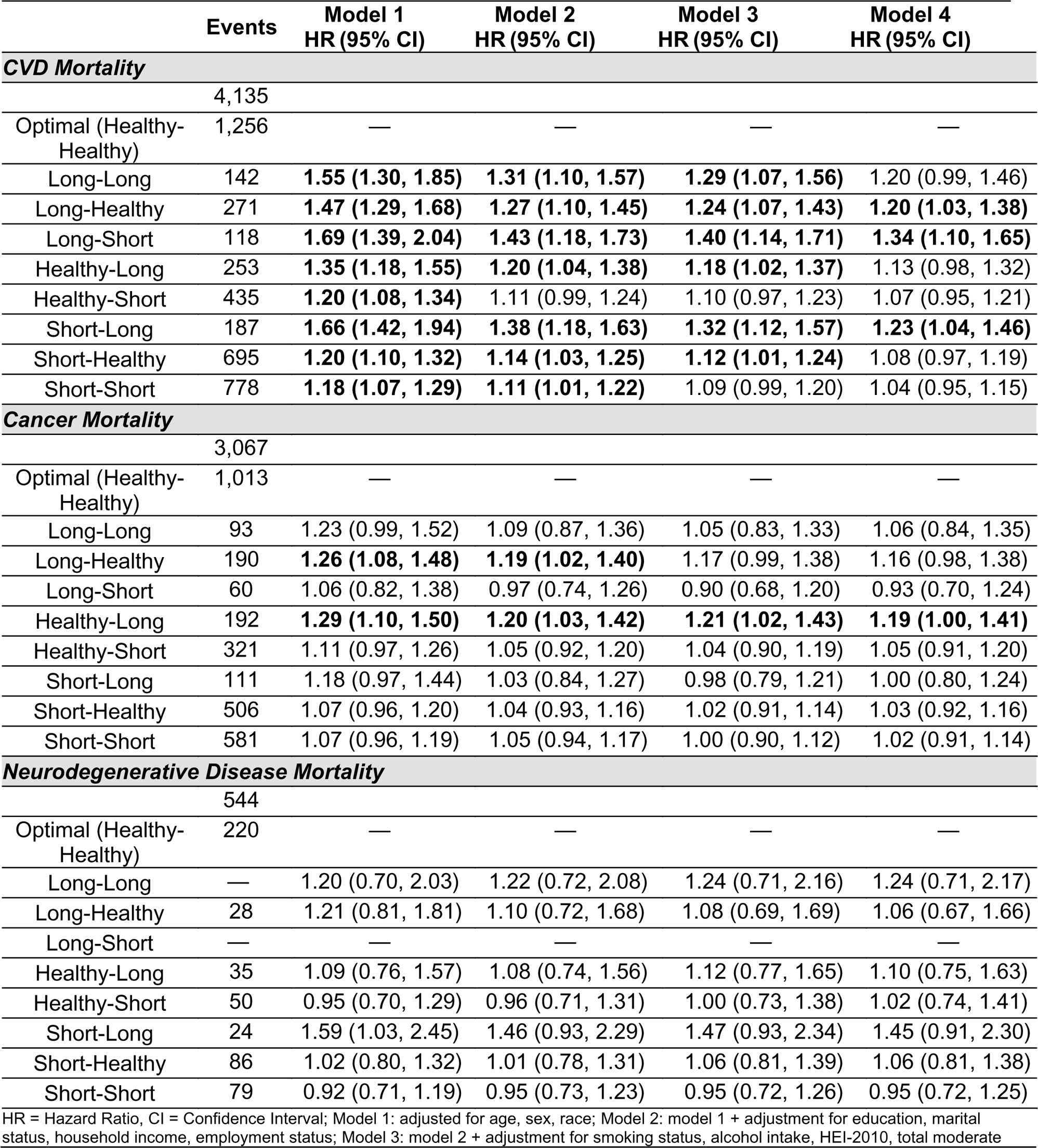
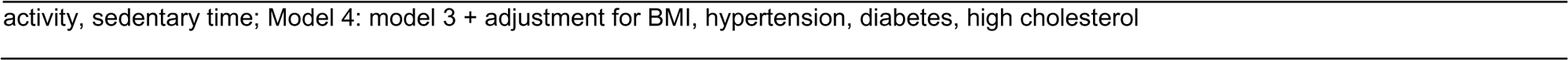
Sleep trajectories and Cause-specific Mortality in the SCCS (N=46,928)

|  | Events | Model 1<br>HR (95% CI) | Model 2<br>HR (95% CI) | Model 3<br>HR (95% CI) | Model 4<br>HR (95% CI) |
| --- | --- | --- | --- | --- | --- |
| **CVD Mortality** |  |  |  |  |  |
|  | 4,135 |  |  |  |  |
| Optimal (Healthy-Healthy) | 1,256 | — | — | — | — |
| Long-Long | 142 | **1.55 (1.30, 1.85)** | **1.31 (1.10, 1.57)** | **1.29 (1.07, 1.56)** | 1.20 (0.99, 1.46) |
| Long-Healthy | 271 | **1.47 (1.29, 1.68)** | **1.27 (1.10, 1.45)** | **1.24 (1.07, 1.43)** | **1.20 (1.03, 1.38)** |
| Long-Short | 118 | **1.69 (1.39, 2.04)** | **1.43 (1.18, 1.73)** | **1.40 (1.14, 1.71)** | **1.34 (1.10, 1.65)** |
| Healthy-Long | 253 | **1.35 (1.18, 1.55)** | **1.20 (1.04, 1.38)** | **1.18 (1.02, 1.37)** | 1.13 (0.98, 1.32) |
| Healthy-Short | 435 | **1.20 (1.08, 1.34)** | 1.11 (0.99, 1.24) | 1.10 (0.97, 1.23) | 1.07 (0.95, 1.21) |
| Short-Long | 187 | **1.66 (1.42, 1.94)** | **1.38 (1.18, 1.63)** | **1.32 (1.12, 1.57)** | **1.23 (1.04, 1.46)** |
| Short-Healthy | 695 | **1.20 (1.10, 1.32)** | **1.14 (1.03, 1.25)** | **1.12 (1.01, 1.24)** | 1.08 (0.97, 1.19) |
| Short-Short | 778 | **1.18 (1.07, 1.29)** | **1.11 (1.01, 1.22)** | 1.09 (0.99, 1.20) | 1.04 (0.95, 1.15) |
| **Cancer Mortality** |  |  |  |  |  |
|  | 3,067 |  |  |  |  |
| Optimal (Healthy-Healthy) | 1,013 | — | — | — | — |
| Long-Long | 93 | 1.23 (0.99, 1.52) | 1.09 (0.87, 1.36) | 1.05 (0.83, 1.33) | 1.06 (0.84, 1.35) |
| Long-Healthy | 190 | **1.26 (1.08, 1.48)** | **1.19 (1.02, 1.40)** | 1.17 (0.99, 1.38) | 1.16 (0.98, 1.38) |
| Long-Short | 60 | 1.06 (0.82, 1.38) | 0.97 (0.74, 1.26) | 0.90 (0.68, 1.20) | 0.93 (0.70, 1.24) |
| Healthy-Long | 192 | **1.29 (1.10, 1.50)** | **1.20 (1.03, 1.42)** | **1.21 (1.02, 1.43)** | **1.19 (1.00, 1.41)** |
| Healthy-Short | 321 | 1.11 (0.97, 1.26) | 1.05 (0.92, 1.20) | 1.04 (0.90, 1.19) | 1.05 (0.91, 1.20) |
| Short-Long | 111 | 1.18 (0.97, 1.44) | 1.03 (0.84, 1.27) | 0.98 (0.79, 1.21) | 1.00 (0.80, 1.24) |
| Short-Healthy | 506 | 1.07 (0.96, 1.20) | 1.04 (0.93, 1.16) | 1.02 (0.91, 1.14) | 1.03 (0.92, 1.16) |
| Short-Short | 581 | 1.07 (0.96, 1.19) | 1.05 (0.94, 1.17) | 1.00 (0.90, 1.12) | 1.02 (0.91, 1.14) |
| **Neurodegenerative Disease Mortality** |  |  |  |  |  |
|  | 544 |  |  |  |  |
| Optimal (Healthy-Healthy) | 220 | — | — | — | — |
| Long-Long | — | 1.20 (0.70, 2.03) | 1.22 (0.72, 2.08) | 1.24 (0.71, 2.16) | 1.24 (0.71, 2.17) |
| Long-Healthy | 28 | 1.21 (0.81, 1.81) | 1.10 (0.72, 1.68) | 1.08 (0.69, 1.69) | 1.06 (0.67, 1.66) |
| Long-Short | — | — | — | — | — |
| Healthy-Long | 35 | 1.09 (0.76, 1.57) | 1.08 (0.74, 1.56) | 1.12 (0.77, 1.65) | 1.10 (0.75, 1.63) |
| Healthy-Short | 50 | 0.95 (0.70, 1.29) | 0.96 (0.71, 1.31) | 1.00 (0.73, 1.38) | 1.02 (0.74, 1.41) |
| Short-Long | 24 | 1.59 (1.03, 2.45) | 1.46 (0.93, 2.29) | 1.47 (0.93, 2.34) | 1.45 (0.91, 2.30) |
| Short-Healthy | 86 | 1.02 (0.80, 1.32) | 1.01 (0.78, 1.31) | 1.06 (0.81, 1.39) | 1.06 (0.81, 1.38) |
| Short-Short | 79 | 0.92 (0.71, 1.19) | 0.95 (0.73, 1.23) | 0.95 (0.72, 1.26) | 0.95 (0.72, 1.25) |

HR = Hazard Ratio, CI = Confidence Interval; Model 1: adjusted for age, sex, race; Model 2: model 1 + adjustment for education, marital status, household income, employment status; Model 3: model 2 + adjustment for smoking status, alcohol intake, HEI-2010, total moderate
activity, sedentary time; Model 4: model 3 + adjustment for BMI, hypertension, diabetes, high cholesterol

In models adjusted for demographic and socioeconomic characteristics, health behaviors, and comorbid conditions the only adverse association observed with cancer mortality was among those participants in the healthy-long sleep trajectory (HR:1.19; CI:1.00, 1.41). While there were no significant associations observed between any of the suboptimal sleep duration trajectories and neurodegenerative disease mortality, however an adverse association was observed for the short-long trajectory that stood out from the other trajectories (in fully adjusted model: HR: 1.45; CI: 0.91, 2.30).

In sensitivity analyses, we observed evidence of effect modification by race and SES indicators. The association between sleep trajectory and all-cause mortality significantly differed among White and Black participants in the healthy-long, healthy-short, long-healthy, short-healthy, short-long, and short-short trajectory categories (all p values <0.05), with greater risk observed among the White participants in those sleep trajectory categories (**Figure 2**). The association between sleep trajectory and all-cause mortality also significantly differed by household income status in the healthy-long, healthy-short, long-healthy, long-long, short-healthy, and short-long trajectory categories (all p values <0.05), with greater mortality risk observed among participants with a household income ≥$15,000 compared to those with a household income <$15,000 (**Figure 3**). Excluding deaths within the first 2 years of follow-up did not substantially impact the results (**Supplemental Table 2 and Table 3**). Excluding participants with a prevalent MI at baseline or follow-up survey did not materially impact observed associations between suboptimal sleep trajectories and CVD mortality (**Supplemental Table 4**). When MICE was employed to account for missing data, associations persisted for all suboptimal sleep trajectories with HRs for all-cause mortality ranging from 7% for the short-short trajectory (HR:1.07; CI: 1.02, 1.12) to 37% for the short-long trajectory (HR: 1.37, CI: 1.25, 1.49) (**Supplemental Table 5**).

**Figure 2.**
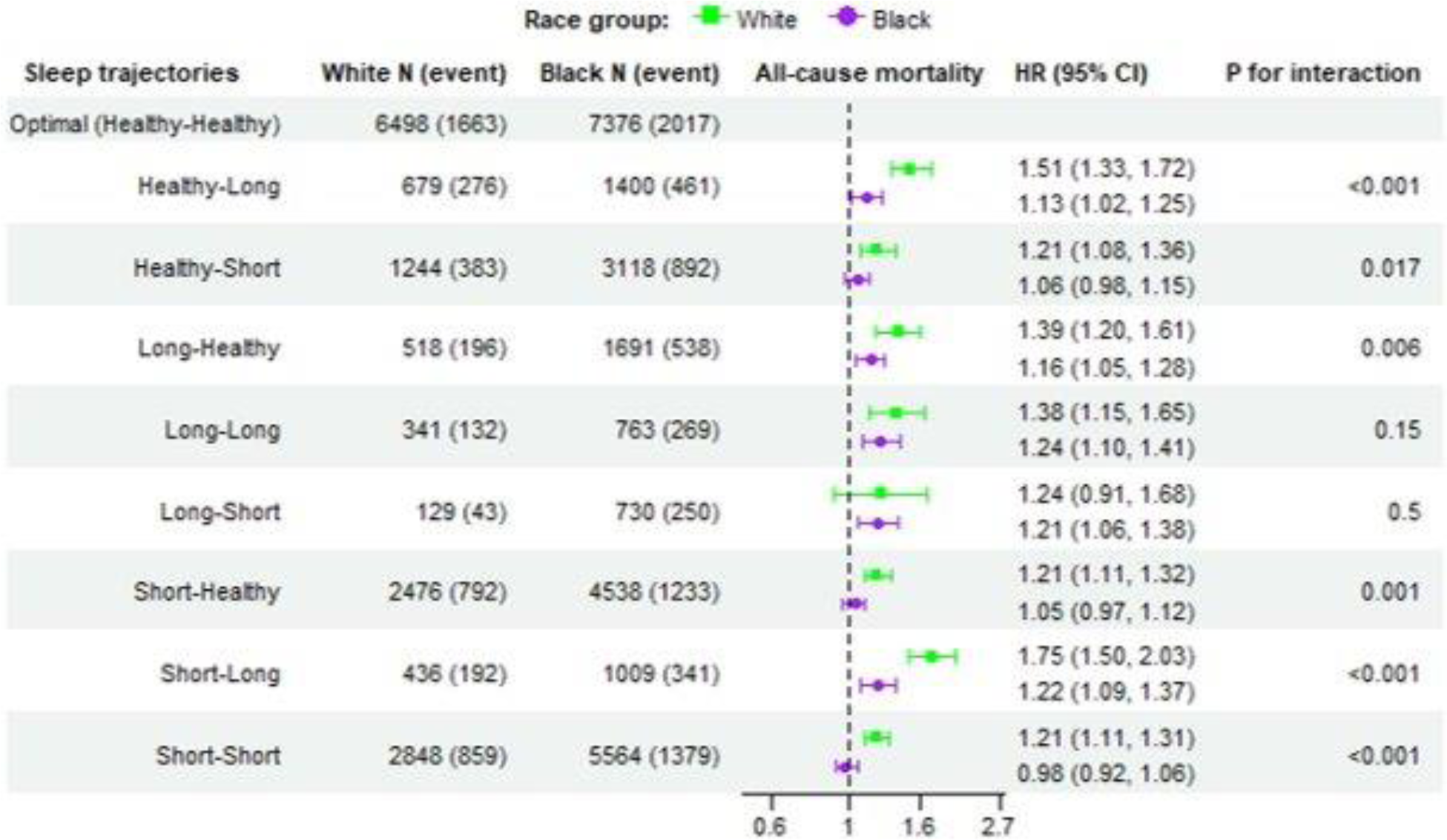
Race-stratified associations between sleep trajectories and all-cause mortality. Adjusted for age, sex, education, marital status, household income, employment status, smoking status, alcohol intake, HEI-2010, total moderate activity, sedentary time.

**Figure 3.**
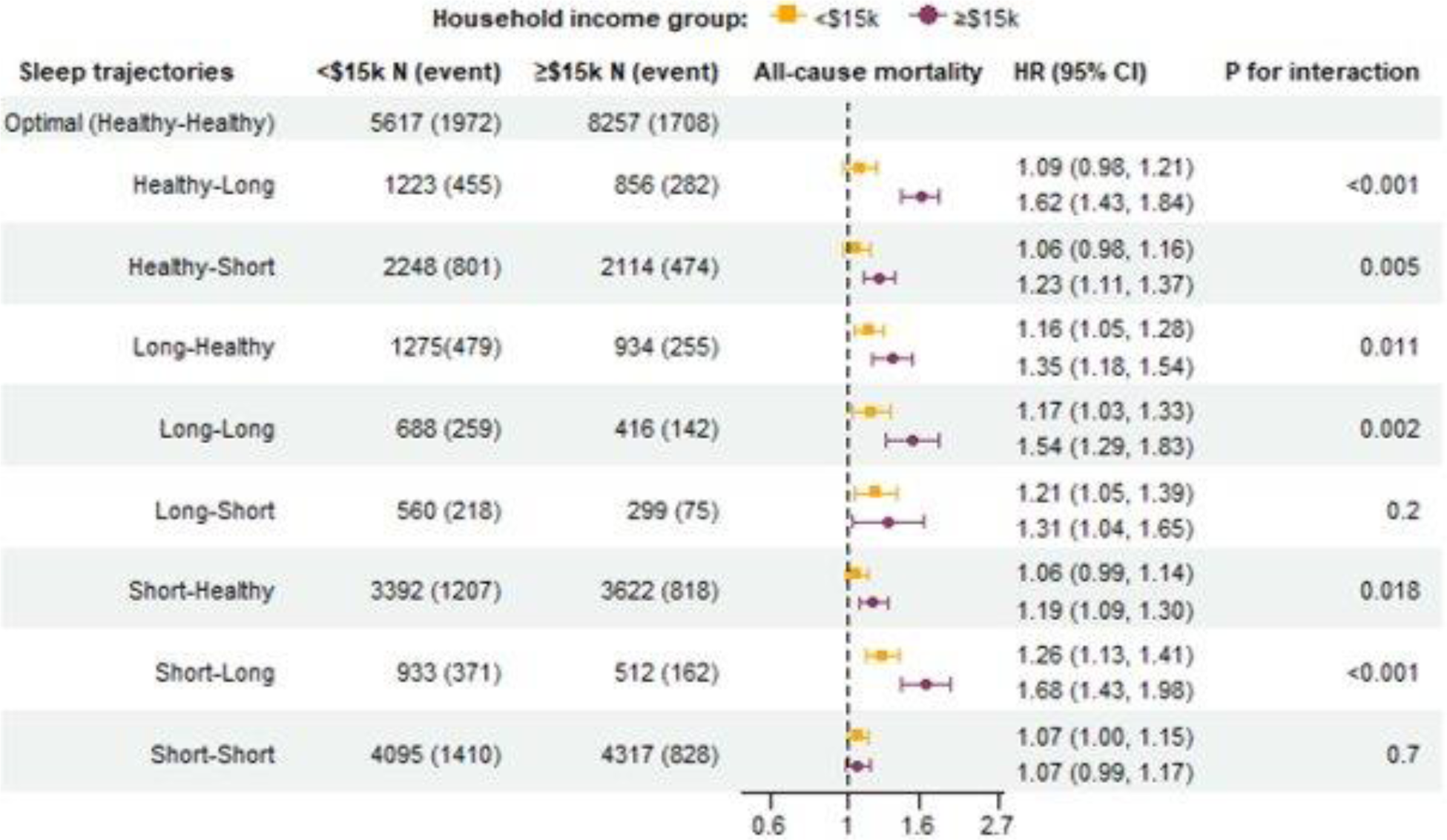
Income-stratified associations between sleep trajectories and all-cause mortality. Adjusted for age, sex, education, marital status, employment status, smoking status, alcohol intake, HEI-2010, total moderate activity, sedentary time.

## Discussion

In a large cohort of low-income middle-aged adults with repeated measures of self-reported sleep duration, we examined the association between sleep duration trajectory with all-cause and cause-specific mortality. Compared to participants who maintained an optimal healthy sleep duration between 7-9 hours over a five-year period, a substantially greater risk of all-cause mortality was observed among individuals in all other sub-optimal sleep trajectory categories. The greatest overall and CVD-specific mortality risk was observed among participants with short-long and long-short sleep trajectories, providing new evidence that irregular sleep patterns over time may increase mortality risk. In race, SES, and sex stratified analyses, the associations were stronger among White adults compared to Black adults and among higher income adults compared to lower-income adults, but there were no differences observed by sex. By examining sleep duration trajectories and all-cause mortality in a large representative cohort of low-income Black and White adults, we are contributing unique and valuable new information on well-documented race and SES sleep disparities.

Our study findings are in line with numerous studies that support the current sleep duration recommendations for 7-9 hours per night for optimal health.^1,2^ However, prior consistent evidence connecting both short and long sleep durations with a wide range of adverse outcomes, particularly cardiovascular risk,^9,11–13,22^ comes primarily from studies in which sleep duration was only assessed at one time point. It is not well understood how habitual sleep patterns and sleep durations change over time within individuals,^23^ and relatively little is known about the association between long-term sleep duration trajectories and mortality risk. Leveraging repeated measures of sleep duration with an average of 4.5 years between timepoints, our study adds new evidence to demonstrate the importance of maintaining a healthy sleep duration (between 7-9 hours) over time in adulthood. In our cohort of over 46,000 adults, only 1/3 of participants reported maintaining a healthy sleep duration from study baseline to follow-up. We were surprised to find nearly 2/3 of participants had irregular or suboptimal sleep duration trajectories over the 4.5-year period. These participants within the irregular or suboptimal trajectory categories were more likely to be Black of African American which is consistent with previous evidence of sleep disparities across racial-ethnic groups.^7,24,25^ In a recent study conducted in the Kailuan cohort among 52,599 Chinese adults, participants reported sleep durations 3 times over a 4-year period.^26^ Approximately 76% of the sample maintained optimal sleep durations between 7-9 hours.^26^ The Kailuan cohort is similar in age to our study cohort, however differences between the studies’ sleep trajectories may be due to the added measurement timepoint, the shorter length of follow-up, or our study being conducted in the US and documented cultural differences in sleep habits between the US and China.^27,28^

We found a consistent 9 to 40% greater risk of all-cause and CVD mortality for participants with suboptimal sleep duration trajectories compared to those with an optimal sleep trajectory. Observed associations are consistent in direction and magnitude with those reported in the Kailuan cohort where suboptimal sleep trajectories were significantly associated with increased risk of CVD events and all-cause mortality (HR for mortality: 1.34 for normal-short trajectory, 1.50 for short-short trajectory).^26^ Our findings are also consistent with those reported in the Sleep Heart Health Study, in which investigators identified sleep duration trajectories based on self-reported sleep duration at baseline and 10 years later in a cohort of predominately White older adults who were on average 63 years at baseline.^29^ After adjustment for demographics, health behaviors, and prevalent CVD risk factors, the short-long (HR: 1.74; 95% CI 1.10, 2.76), healthy-long (HR: 1.63; 95% CI 1.26, 2.13), and long-short (HR:1.99; 95% CI 1.03, 4.05) trajectories were associated with a nearly 2-fold greater risk of all-cause mortality.^29^ These associations persisted after adjustment for objectively-measured sleep disturbed breathing, an important and relevant CVD risk factor. Similarly, in our analyses we adjusted for pertinent CVD risk factors and found associations persisted across all sub-optimal sleep trajectories for all-cause mortality and almost all trajectories for CVD mortality, with the greatest risk observed among the short-long and the long-short trajectories. Our findings from a large cohort of adults at mid-life are an important new contribution to the literature that suggest that maintaining optimal sleep duration throughout midlife may play an important role in reducing mortality risk.

A strength of this study was the appropriate statistical power to investigate subgroup differences. We observed significant variations in the associations by race and SES indicators, but not sex. In almost all of the sleep trajectory categories we observed a greater magnitude of association between suboptimal sleep trajectory and all-cause mortality among White participants as compared to Black participants, and among individuals with greater household incomes as compared to those with lower household incomes. These findings were surprising considering SCCS participants with an optimal sleep duration trajectory were more likely to identify as White and the well-documented sleep disparities and greater cardiovascular risk observed among both Black and African American adults and in low-income communities.^24,30,31^ Although the Black and low-SES participants in the SCCS were more likely to have suboptimal sleep trajectories, our results suggest the potential health effects of suboptimal sleep may be weaker in these populations. This may be due in part to the presence of more risk factors (e.g. smoking) in these populations, which may confound or attenuate the effect of sleep duration trajectory. Regardless, it is important to note that an association between suboptimal sleep trajectory and risk of mortality was still observed among the Black and low-income subgroups, suggesting that sleep duration trajectory is still an important risk factor that warrants attention. While hypothesis generating, these results suggest that there may be other factors, beyond the social constructs of race and income, such as physical and social environments or occupational factors, that are not being captured and may negatively affect sleep health and ultimately contribute to mortality.^31^ These results also highlight the importance of examining sleep health in new samples that offer more diverse representation not only in terms of race, but also in geography and SES.

There are several additional study findings that are worth noting. Despite our hypotheses, we only observed an association between the healthy-long sleep trajectory and cancer mortality. Previous findings on sleep duration and cancer mortality have been mixed, with some inconsistent evidence suggesting that long sleepers may be at a greater mortality risk. In a meta-analysis of 32 studies, Stone et al. reported for long sleepers a pooled HR of 1.09 (95% CI 1.04–1.13) for all-cancer mortality, indicating that compared to recommended sleep duration of 7-9 hours, those sleeping longer were at a 9% greater cancer mortality risk.^32^ In a mendelian randomization study conducted in the UK Biobank Cohort, Tian et al reported a linear association between long sleep duration and all-cause mortality in cancer patients (OR, 5.56, 95% CI: 3.15-9.82, p = 3.42E-09), suggesting that as sleep duration increases so does risk of mortality in patients with cancer.^33^ In our study, the greater cancer mortality risk observed among participants in the healthy-long trajectory is consistent with these previous findings. In sensitivity analyses, after excluding deaths within the first 2 years of follow-up, the long-healthy trajectory, but not the healthy-long, trajectory was associated with cancer mortality in fully adjusted models. More research is needed to better understand the connection between long sleep duration and cancer mortality risk.

We observed no significant associations between any suboptimal sleep trajectory and risk of neurodegenerative disease mortality. These null findings may be due to the small number of neurodegenerative disease mortality events. There was a suggestion of an increased risk of neurodegenerative disease mortality among those with a short-long sleep trajectory that may reflect age-related changes in sleep patterns and is worth further exploration. The potential for underreporting of Alzheimer’s disease and Parkinson’s disease on death certificates has been documented,^34,35^ and this could attenuate observed associations between sleep trajectories and mortality.

Emerging evidence suggests sleep regularity is an important sleep behavior to target to reduce cardiovascular risk.^36,37^ To date, sleep regularity has traditionally been defined as variation in sleep durations across a one-week period. In our cohort, sleep duration was measured at 2 timepoints, approximately 4.5 years apart. Participants who went from either a short or long sleep duration at enrollment to a healthy sleep duration at follow up were still at a greater risk for all-cause and CVD mortality compared to participants who maintained optimal healthy sleep durations across the 4.5-year period. These results support the lasting impact of suboptimal sleep, and suggest that beyond achieving a healthy sleep duration, maintaining regular sleep patterns over longer periods of time is another potentially relevant risk reducing health behavior. Our findings provide novel evidence that long-term sleep duration irregularity may be adversely associated with CVD mortality, independent of socioeconomic factors, health behaviors, and CVD risk factors.

Our study has several strengths. We leveraged data from a large prospective cohort of low-income Black and White Americans who have been historically underrepresented in other US cohorts. Participants self-reported their sleep duration at two timepoints over a 4.5-year period and overall and cause-specific mortality data for all participants were obtained from linkage with the National Death Index. Our study also has some limitations. With our self-reported sleep measure, we were only able to capture one measure of sleep health (duration) and were not able to account for other potentially relevant dimensions of sleep health or sleep disorders, such as sleep apnea or insomnia, which may be important confounders. It is common for adults to overestimate their sleep duration,^38^ and we may have misclassified participants which likely would bias our associations towards the null. Finally, our study is observational and therefore we are unable to rule out residual confounding or establish causality.

In conclusion, our study connected suboptimal long-term sleep duration trajectories to greater risk of all-cause and CVD-specific mortality in a large cohort of predominantly low-income Black and White Americans. In stratified analyses, we observed the association between suboptimal sleep trajectory and all-cause mortality were stronger among the White participants compared to the Black participants in our study and among those with a great household income as compared to the lower household income. The associations observed were independent of relevant sociodemographic, behavioral, and CVD risk factors. Our findings highlight the importance of maintaining healthy and regular sleep durations throughout mid-life.

## Data Availability

All SCCS Data, including data supporting this study are available upon application at www.southerncommunitystudy.org. Access to the data is subject to approval and a data sharing agreement.

## STROBE Statement—checklist of items that should be included in reports of observational studies

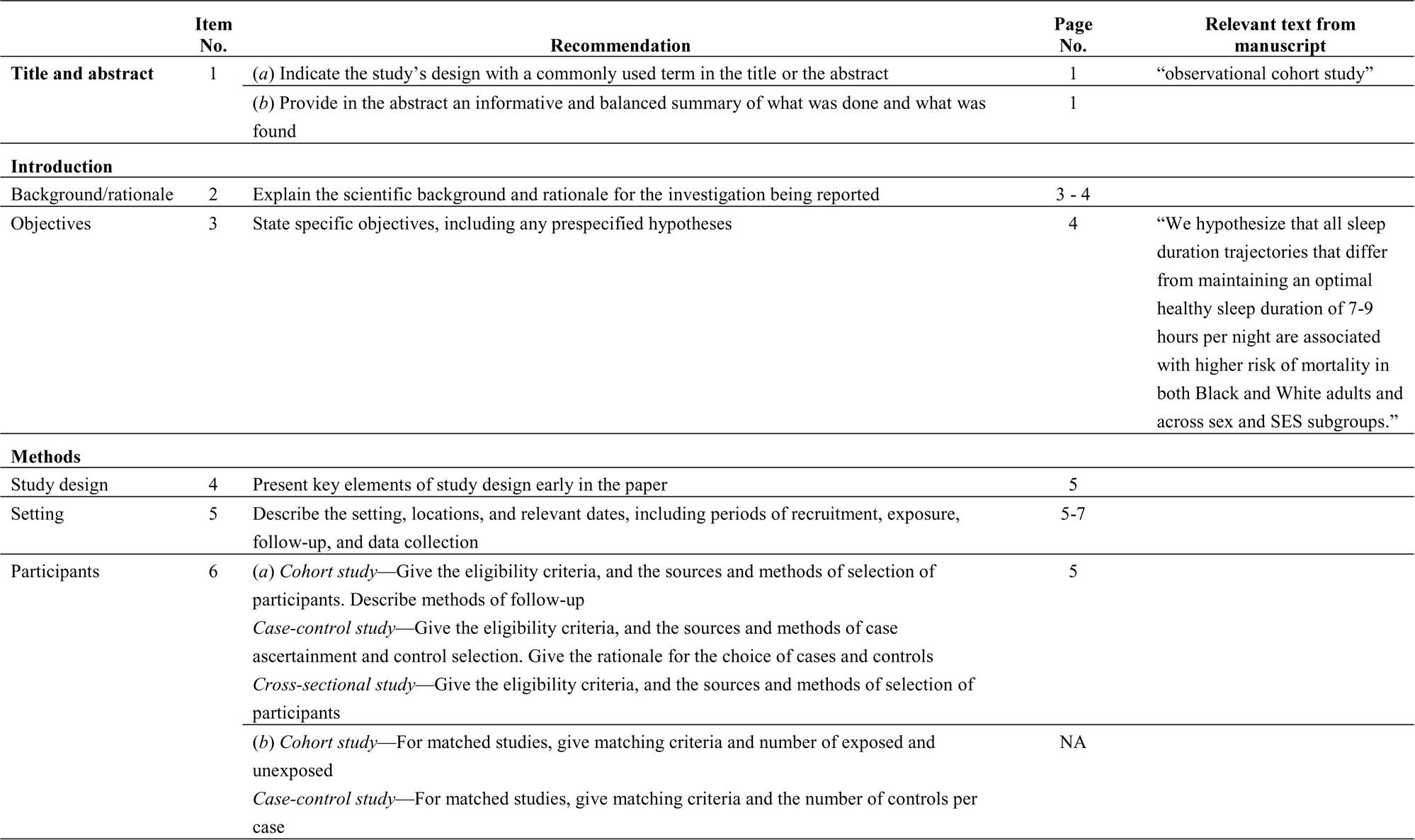
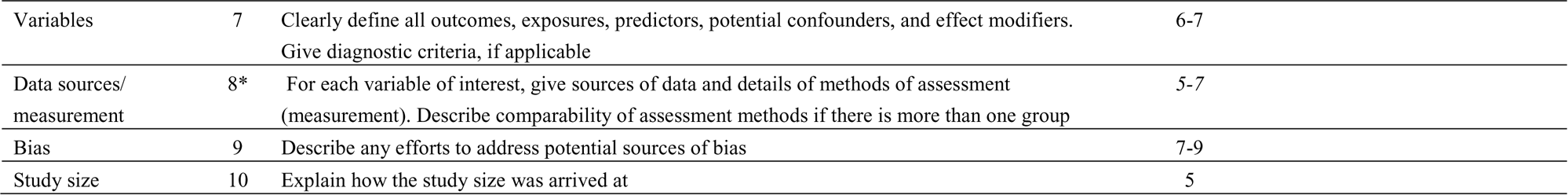
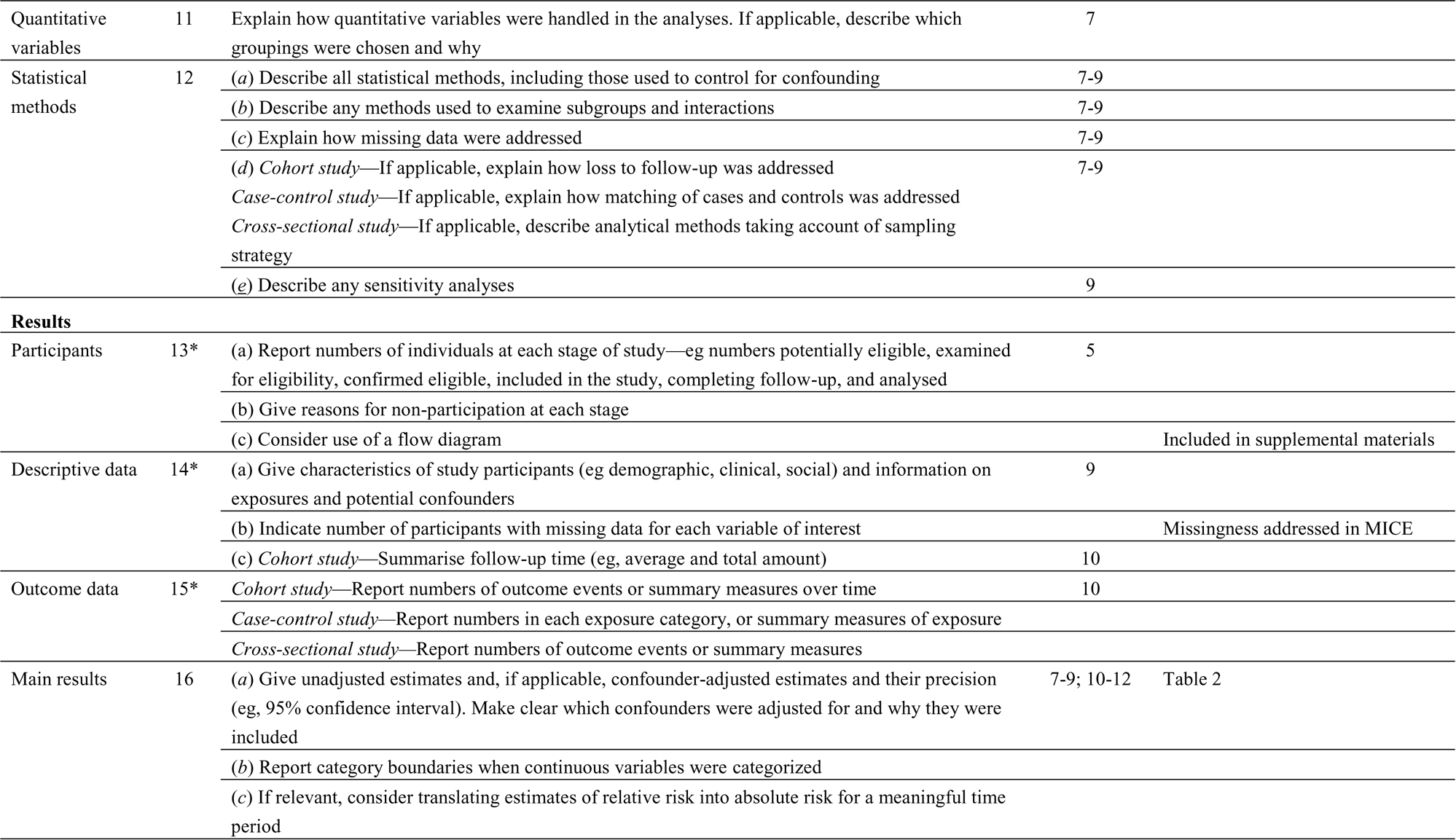
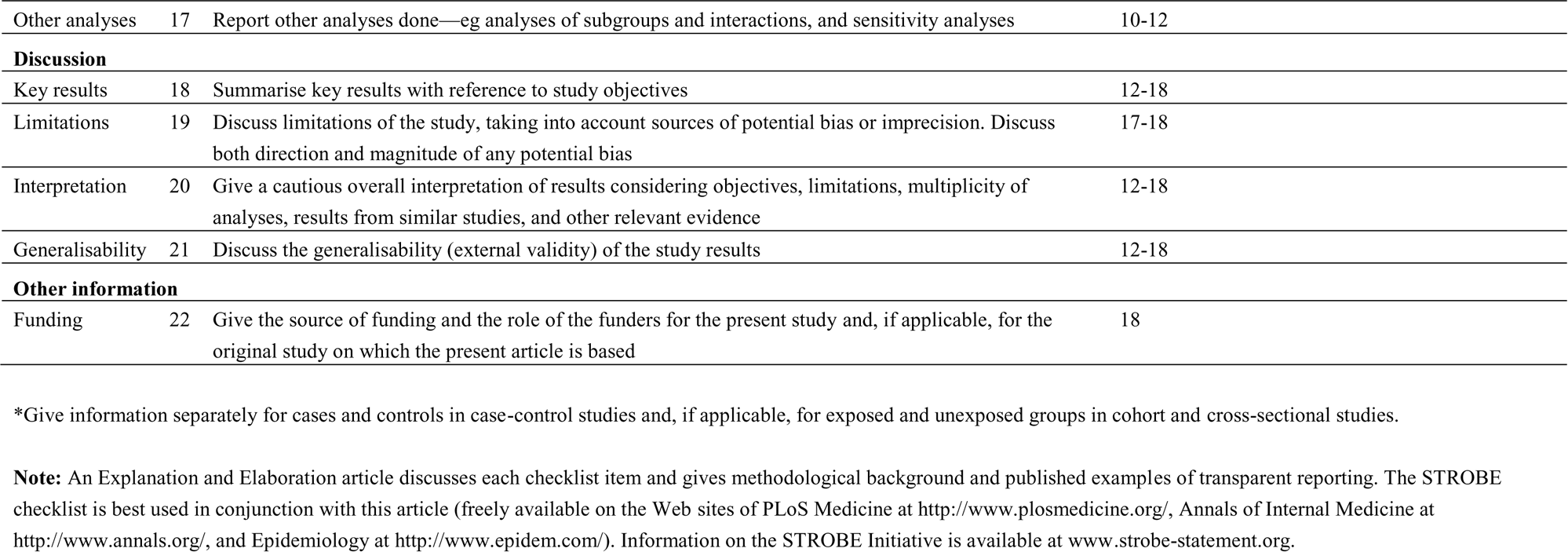

|  | Item No. | Recommendation | Page No. | Relevant text from manuscript |
| --- | --- | --- | --- | --- |
| Title and abstract | 1 | (a) Indicate the study’s design with a commonly used term in the title or the abstract | 1 | “observational cohort study” |
|  |  | (b) Provide in the abstract an informative and balanced summary of what was done and what was found | 1 |  |
| Introduction |  |  |  |  |
| Background/rationale | 2 | Explain the scientific background and rationale for the investigation being reported | 3 - 4 |  |
| Objectives | 3 | State specific objectives, including any prespecified hypotheses | 4 | “We hypothesize that all sleep duration trajectories that differ from maintaining an optimal healthy sleep duration of 7-9 hours per night are associated with higher risk of mortality in both Black and White adults and across sex and SES subgroups.” |
| Methods |  |  |  |  |
| Study design | 4 | Present key elements of study design early in the paper | 5 |  |
| Setting | 5 | Describe the setting, locations, and relevant dates, including periods of recruitment, exposure, follow-up, and data collection | 5-7 |  |
| Participants | 6 | (a) Cohort study—Give the eligibility criteria, and the sources and methods of selection of participants. Describe methods of follow-up<br>Case-control study—Give the eligibility criteria, and the sources and methods of case ascertainment and control selection. Give the rationale for the choice of cases and controls<br>Cross-sectional study—Give the eligibility criteria, and the sources and methods of selection of participants | 5 |  |
|  |  | (b) Cohort study—For matched studies, give matching criteria and number of exposed and unexposed<br>Case-control study—For matched studies, give matching criteria and the number of controls per case | NA |  |

|  |  |  |  |
| --- | --- | --- | --- |
| Variables | 7 | Clearly define all outcomes, exposures, predictors, potential confounders, and effect modifiers.<br>Give diagnostic criteria, if applicable | 6-7 |
| Data sources/<br>measurement | 8* | For each variable of interest, give sources of data and details of methods of assessment (measurement). Describe comparability of assessment methods if there is more than one group | 5-7 |
| Bias | 9 | Describe any efforts to address potential sources of bias | 7-9 |
| Study size | 10 | Explain how the study size was arrived at | 5 |
| Quantitative variables | 11 | Explain how quantitative variables were handled in the analyses. If applicable, describe which groupings were chosen and why | 7 |  |
| Statistical methods | 12 | (a) Describe all statistical methods, including those used to control for confounding | 7-9 |  |
|  |  | (b) Describe any methods used to examine subgroups and interactions | 7-9 |  |
|  |  | (c) Explain how missing data were addressed | 7-9 |  |
|  |  | (d) *Cohort study*—If applicable, explain how loss to follow-up was addressed | 7-9 |  |
|  |  | *Case-control study*—If applicable, explain how matching of cases and controls was addressed |  |  |
|  |  | *Cross-sectional study*—If applicable, describe analytical methods taking account of sampling strategy |  |  |
|  |  | (e) Describe any sensitivity analyses | 9 |  |
| **Results** |  |  |  |  |
| Participants | 13* | (a) Report numbers of individuals at each stage of study—eg numbers potentially eligible, examined for eligibility, confirmed eligible, included in the study, completing follow-up, and analysed | 5 |  |
|  |  | (b) Give reasons for non-participation at each stage |  |  |
|  |  | (c) Consider use of a flow diagram |  | Included in supplemental materials |
| Descriptive data | 14* | (a) Give characteristics of study participants (eg demographic, clinical, social) and information on exposures and potential confounders | 9 |  |
|  |  | (b) Indicate number of participants with missing data for each variable of interest |  | Missingness addressed in MICE |
|  |  | (c) *Cohort study*—Summarise follow-up time (eg, average and total amount) | 10 |  |
| Outcome data | 15* | *Cohort study*—Report numbers of outcome events or summary measures over time | 10 |  |
|  |  | *Case-control study*—Report numbers in each exposure category, or summary measures of exposure |  |  |
|  |  | *Cross-sectional study*—Report numbers of outcome events or summary measures |  |  |
| Main results | 16 | (a) Give unadjusted estimates and, if applicable, confounder-adjusted estimates and their precision (eg, 95% confidence interval). Make clear which confounders were adjusted for and why they were included | 7-9; 10-12 | Table 2 |
|  |  | (b) Report category boundaries when continuous variables were categorized |  |  |
|  |  | (c) If relevant, consider translating estimates of relative risk into absolute risk for a meaningful time period |  |  |

|  |  |  |  |
| --- | --- | --- | --- |
| Other analyses | 17 | Report other analyses done—eg analyses of subgroups and interactions, and sensitivity analyses | 10-12 |
| **Discussion** |  |  |  |
| Key results | 18 | Summarise key results with reference to study objectives | 12-18 |
| Limitations | 19 | Discuss limitations of the study, taking into account sources of potential bias or imprecision. Discuss both direction and magnitude of any potential bias | 17-18 |
| Interpretation | 20 | Give a cautious overall interpretation of results considering objectives, limitations, multiplicity of analyses, results from similar studies, and other relevant evidence | 12-18 |
| Generalisability | 21 | Discuss the generalisability (external validity) of the study results | 12-18 |
| **Other information** |  |  |  |
| Funding | 22 | Give the source of funding and the role of the funders for the present study and, if applicable, for the original study on which the present article is based | 18 |

\*Give information separately for cases and controls in case-control studies and, if applicable, for exposed and unexposed groups in cohort and cross-sectional studies.
**Note:** An Explanation and Elaboration article discusses each checklist item and gives methodological background and published examples of transparent reporting. The STROBE checklist is best used in conjunction with this article (freely available on the Web sites of PLoS Medicine at http://www.plosmedicine.org/, Annals of Internal Medicine at http://www.annals.org/, and Epidemiology at http://www.epidem.com/). Information on the STROBE Initiative is available at www.strobe-statement.org.

